# Evaluation of SARS-CoV-2 rapid antigen diagnostic tests for saliva samples

**DOI:** 10.1101/2021.05.14.21257100

**Authors:** Marie Hagbom, Noelia Carmona-Vicente, Sumit Sharma, Henrik Olsson, Mikael Jämtberg, Åsa Nilsdotter-Augustinsson, Johanna Sjöwall, Johan Nordgren

## Abstract

**Background:** The COVID-19 pandemic has highlighted the need for rapid, cost effective and easy-to-use diagnostic tools for SARS-CoV-2 rapid antigen detection (RAD) for use in point of care settings or as self-tests, to limit disease transmission. Using saliva samples would further greatly facilitate sample collection, diagnostic feasibility, and mass screening.

**Objective:** We tested two rapid antigen immunochromatographic tests designed for detection of SARS-CoV-2 in saliva: Rapid Response™ COVID-19 Antigen Rapid Test Cassette for oral fluids (Rapid Response) and DIAGNOS™ COVID-19 Antigen Saliva Test (DIAGNOS). Evaluation of detection limit was performed with purified SARS-CoV-2 nucleocapsid protein and titrated live SARS-CoV-2 virus and compared to Abbott Panbio™ COVID-19 Ag Rapid Test (Panbio) designed for nasopharyngeal samples. Sensitivity and specificity were further evaluated on RT-qPCR positive and negative saliva samples from individuals hospitalized with COVID-19 (n=34); and asymptomatic health care personnel (n=20).

**Results:** The limit of detection of the saliva test from DIAGNOS was comparable with the Panbio test and showed higher sensitivity than Rapid Response for both nucleocapsid protein and diluted live viruses. DIAGNOS and Rapid Response further detected seven (47%) and five (33%), respectively, of the 15 RT-qPCR positive saliva samples in individuals hospitalized with COVID-19. Of the 39 RT-qPCR negative samples, all were negative with both tests (specificity 100%; 95% c.i. 0.91-1.00). Only one of the RT-qPCR positive saliva samples (Ct 21.6) contained infectious virus as determined by cell culture and was also positive using the saliva RADs.

**Conclusion:** The results show that the DIAGNOS test exhibit a similar limit of detection as the Panbio RAD and may be an important and easy-to-use saliva RAD complement to detect infectious individuals.

## Introduction

There is an urgent need for rapid and easy to use diagnostics for SARS-CoV-2 to limit disease transmission during the ongoing pandemic. The diagnostic gold standard, reverse transcription real-time PCR (RT-qPCR) have a high specificity and sensitivity, but have limitations with regards to time, cost, and logistics (1, 2).

To overcome these issues several rapid antigen diagnostic tests (RADs) have rapidly been developed for diagnosis of SARS-CoV-2. There are now a multitude of such RADs available (3). At least a few have demonstrated good sensitivity and specificity in comparison to RT-qPCR (3-6). Moreover, previous studies have shown that RADs can exhibit high sensitivity in detecting samples containing infectious virus, indicating a high sensitivity to detect contagious individuals (7, 8). The vast majority of RADs today are designed for nasopharyngeal samples (3-6). Using saliva instead of nasopharyngeal swabs has several advantages. It is a noninvasive technique, easy to self-collect with little discomfort, does not require specialized health care personnel and thus reduced risk for the user (9). Today, there are few RADs designed for saliva available that have been thoroughly validated.

In this study, we have evaluated saliva RADs from two suppliers: Rapid Response™ COVID-19 Antigen Rapid Test Cassette for oral fluids (Rapid Response) and DIAGNOS™ COVID-19 Antigen Saliva Test (DIAGNOS). Both RADs are immunochromatographic assays detecting the SARS-CoV-2 nucleocapsid protein in saliva without any specialized instruments. The tests were first evaluated using SARS-CoV-2 nucleocapsid protein and live SARS-CoV-2 virus titrated in saliva or test kit buffer and compared to Abbott Panbio™ COVID-19 Ag Rapid Test (Panbio) designed for nasopharyngeal samples. Finally, the tests were evaluated on saliva samples from individuals hospitalized with COVID-19 and asymptomatic health care personnel and compared to RT-qPCR. We further correlated the sensitivity of the RADs with regards RT-qPCR Ct-values and infectivity in cell culture.

## Material and Methods

### Clinical samples and rapid antigen saliva tests

Saliva samples were obtained from COVID-19 hospitalized individuals in the COVID-19 cohort of Vrinnevi hospital, Norrköping, Sweden. Saliva samples were taken by a nurse and after being transported to the laboratory the same day, aliquoted and stored at -80°C until analysis. In total 34 saliva samples were used in this study, collected five to 30 days post symptom onset. Saliva samples from asymptomatic healthcare workers (n=20) were also analyzed. The two antigen saliva tests used in the study were Rapid Response™ COVID-19 Antigen Rapid Test Cassette for oral fluids (BTNX, Markham, Canada) and DIAGNOS™ COVID-19 Antigen Saliva Test kit (Nantong Diagnos Biotechnology, Rugao, China).

### Sensitivity testing with recombinant SARS-CoV-2 nucleocapsid protein

To test the sensitivity and detection limit of the nucleocapsid protein (NCp), purified NCp (Nordic Biosite, Sweden, Code: OOEF01087) at a concentration ranging between 500 ng to 5 pg, with 10-fold dilutions was tested with both kits. Each kit buffer was used for making the dilution series. One drop of kit buffer was added, followed by 50 µL of sample containing the protein and two more drops of buffer. Both kits were tested at the same time to make it possible to directly compare the readings. After 15 minutes of incubation, two persons, independently of each other, made the readings which were documented.

### Sensitivity of rapid antigen tests using saliva diluted SARS-CoV-2 virus

Saliva negative for SARS-CoV-2 were used to make dilutions of cell cultured infectious SARS-CoV-2. A 10-fold dilution series starting from 1:10 to 1:1.000.000, were prepared and used to test both RADs according to the respective manufacturers’
s instructions. The same saliva without virus addition was used as negative control.

### Evaluation on saliva collected from patients hospitalized with COVID-19

Samples were tested simultaneously with both antigen tests to enable a direct comparison. According to instructions, 30 µl of saliva + 70 µl of kit buffer were mixed for the Rapid Response™ COVID-19 Antigen Rapid Test Cassette, and 40 µl of saliva + 35 µl of kit buffer for DIAGNOS™ COVID-19 Antigen Saliva Test kit. The prepared sample was added to each test stick in the given volume (100 μL and 75 μL, respectively), at room temperature, according to the manufacturer’s instructions. After 15 minutes of incubation, two persons, independently of each other, read all test sticks to determine positivity or negativity. Positivity was further categorized into three strengths: “+++” where the intensity of the test band was stronger than the control band, “++” where the test band intensity was similar to the control band and “+” where the test band intensity was weaker. There was no difference in observations made by the two persons. Photos were taken for documentation.

### Cell culture to detect infectious virus in saliva

All SARS-CoV-2 PCR positive saliva samples were cultured on Vero E6 cells. Vero E6 cells were cultured in Dulbecco’s Modified Eagle Medium (DMEM) supplemented with 10% fetal calf serum (FCS) and gentamycin. Cells were seeded as monolayer in 48-well plates and at time of confluency they were infected with the saliva sample in a biosafety level 3 (BSL3) laboratory, essentially as described (7). Before infection the cells were washed two times with DMEM supplemented with 2% FCS and gentamycin. 20 μL of saliva samples were diluted (DMEM supplemented with 2% FCS and gentamycin) to a total volume of 350 μL in a 48 well with cells. Samples were blind passaged two times on Vero E6 cells for 3 days each. All cultured saliva samples were tested after passage two for presence of SARS-CoV-2 using the Panbio antigen test and RT-qPCR, to investigate virus replication.

### RNA extraction and in-house RT-qPCR for envelope and RdRp genes

Viral RNA was extracted using QIAmp Viral RNA kit (Qiagen, Hilden, Germany) according to manufactures instructions with the exception that 10 times less sample volume was used, due to need of saliva for both the two RADs as well as cell culture. RNA from the COVID-19 cohort saliva samples (n=54), saliva containing titration of SARS-CoV-2 virus and cultured saliva samples on Vero E6 cells were analyzed using a qPCR for the envelope and RdRp genes. The primers and probes for the envelope gene were E_Sarbeco_P1, E_Sarbeco_F and E_Sarbeco_R from (10). The primers for RdRp were RdRpF: 5’-GTC ATG TGT GGC GGT TCA CT-‘3 and RdRpR 5’-AAA CAC TAT TAG CAT AAG CAG TTG-’3, modified from (11), and probe RdRp_Pi 5’ AGG TGG AAC CTC ATC AGG AGA TGC ‘3 from (11). RT-qPCR was performed on CFX96 (Biorad) using iTaq Universal Probes Supermix (Biorad) with following cycling conditions: reverse transcription at 46°C for 30 min; followed by initial denaturation at 95°C for 3 min, followed by 45 cycles of 95°C for 5 seconds and 56°C for 1 minute (RdRp primers) or 58°C for 30 seconds (envelope primers). All samples were run in duplicates. A sample was considered positive if any of the two RT-qPCR assays were positive. Saliva samples from asymptomatic healthcare workers were only analyzed for the RdRp gene.

### Ethical statement

Saliva sample collection from COVID-19 patients and asymptomatic personnel were a part of a study approved by the Ethical board in Linköping; Dnr: 202002580 and Dnr: 2021-00419.

## Results and discussion

In this study we have evaluated two SARS-CoV-2 RADs designed for use in saliva samples, enabling noninvasive, easy sampling. Firstly, we investigated the detection limit of the assays comparing to Panbio, a widely used RAD for nasopharyngeal samples, using the SARS-CoV-2 NCp protein in a range concentration from 500 ng to 5 pg. The Rapid Response test detected 50 pg, and the DIAGNOS test as well as the Panbio detected 5 pg. These detection limits are in the range of conventional ELISAs (12), which is a standard method of protein detection. We subsequently performed two different sets of virus detection tests, using dilution series of laboratory cultivated SARS-CoV-2 diluted in i) saliva samples, or in ii) test kit buffer, the latter enabling a direct comparison with the Panbio RAD. The DIAGNOS and Rapid Response detected the same virus amount as the Panbio using SARS-CoV-2 diluted in test kit buffer (data not shown), whereas the DIAGNOS test exhibited a lower detection limit for virus diluted in saliva compared to the Rapid Response test (Figure 1).

**Figure 1.**
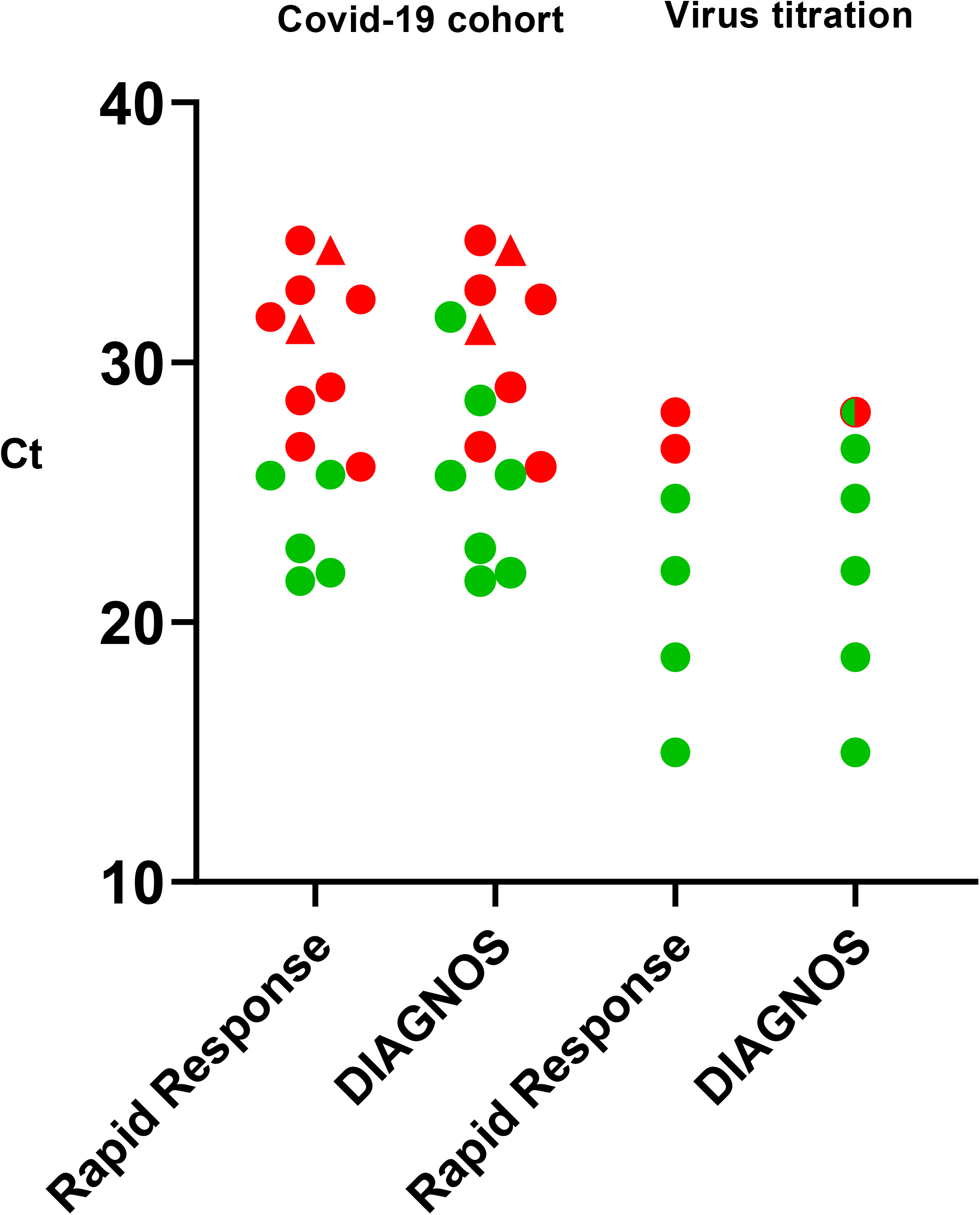
Scatter dot blot indicating positivity (green dot) and negativity (red dot) of the rapid saliva antigen tests in association to RT-qPCR Ct-values for the envelope gene. Two saliva samples were positive only for the RdRp gene (triangle) and are marked with corresponding RdRp Ct values. DIAGNOS™ detected seven and Rapid Response™ five of the 15 RT-qPCR positive samples in the COVID-19 cohort (left panel). Using live SARS-CoV-2 titrated in 10-fold dilutions of saliva (right panel), DIAGNOS test exhibited higher sensitivity compared to the Rapid Response test. Green/red dot indicate weak positivity (+/-).

We subsequently tested the saliva samples collected from patients hospitalized for COVID-19 using the RADs (Table 1). The saliva was collected at various time points post symptom onset, with a median of 10.5 days (range 5-30 days). 15 saliva samples were RT-qPCR positive, and 19 saliva samples were RT-qPCR negative. 13 of the 15 RT-qPCR positive saliva were positive with both envelope and RdRp gene assays, whereas two were only positive with the RdRp assay (Figure 1). In addition to saliva samples of hospitalized patients, saliva samples from asymptomatic health care workers were also included (n=20), all of which were negative by RT-qPCR.

**Table 1.** Characteristics of the COVID-19 cohort in association to SARS-CoV-2 positivity

|  | Presence of viral RNA in saliva |  |  | RAD of RT-qPCR pos saliva |  |  |
| --- | --- | --- | --- | --- | --- | --- |
|  | Saliva samples | RT-qPCR<sup>a</sup> |  | Rapid Response | DIAGNOS | Infectivity<sup>b</sup> |
|  |  | Pos | Neg |  |  |  |
| **COVID-19 hospitalized patients (N)** | 34 | 15/34 | 19/34 | 5/15 | 7/15 | 1/15 |
| **Gender** |  |  |  |  |  |  |
| **Male** | 26 (76.5%) | 12/26 (46.2%) | 14/26 (53.8%) | 5/12 (41.7%) | 6/12 (50.0%) | 1/12 |
| **Female** | 8 (23.5%) | 3/8 (37.5%) | 5/8 (62.5%) | 0/3 (0%) | 1/3 (33.3%) | 0/3 |
| **Median age (yr)** | 57.5 (32-78) | 60.0 (39-78) | 56.0 (32-73) | 60.0 (51-75) | 59.0 (47-75) | 51 |
| **Median DPS** | 10.5 (5-30) | 10.0 (5-30) | 11.0 (7-24) | 6.0 (5-30) | 6.0 (5-30) | 6 |
| **Healthcare workers** | 20 | 0/20 | 20/20 | 0/20 | 0/20 |  |

a) 13 saliva samples were positive for both the envelope and RdRp genes, whereas 2 samples were only positive for the RdRp gene
b) Containing infectious virus as determined by CPE and replication in cell culture
DPS: Days post symptom onset

In total, all RT-qPCR negative saliva samples (n=39), 20 samples from the health care workers and 19 from patients hospitalized with COVID-19, were negative with both tests (specificity 100%; 95% c.i. 0.91-1.00).

Of the 15 RT-qPCR positive samples from hospitalized COVID-19 patients, seven (47%) were positive with the DIAGNOS test and five (33%) with Rapid Response test (Figure 1). As expected, subgroup analysis showed an association between lower Ct-values and RAD positivity (Figure 1). We then proceeded to assess the correlation between infectivity and RT-qPCR results. Only one of the 15 PCR positive (6.7%) saliva samples showed SARS-CoV-2 replication and associated CPE in Vero E6 cells. This sample contained the highest viral load (Ct 21.6) of all the saliva samples in the study. This saliva sample, collected 6 days after onset of symptoms, was positive with both RADs. Although the numbers are limited, these findings are of interest as they indicate that the saliva RADs can have high sensitivity for detecting contagious individuals, as previously reported for nasopharyngeal RADs (7, 8, 13). To note, one sample with a Ct of 21.9 collected at day 5 post symptom onset, did not show replication or associated CPE in cell culture. Sample infectivity, as determined by cell culturing, may not ultimately define whether a person is infectious or not, but detection of infectious virus in cell culture can be a more reliable method than PCR, since PCR does not provide information about virus viability (14, 15).

Several studies have evaluated the use of saliva instead of nasopharyngeal swabs as a clinical specimen for COVID-19 diagnostics. Most studies detecting viral RNA in saliva have been evaluated in symptomatic or hospitalized patients, although a few studies screening saliva in asymptomatic individuals exist as well. The results are heterogenous, with saliva showing lower diagnostic accuracy in some studies, while higher in others (9, 13, 16-18);. with one study reporting a higher concordance to nasopharyngeal samples early after symptom onset (18). In this study, no major differences were observed between RT-qPCR positivity with regards to days post symptom onset, with positive saliva having a median of 10 and a negative saliva a median of 11 days (Table 1). The evaluated saliva RADs, however, were more likely to yield a positive result if the saliva sample was collected early during the infection (median 6 days) (Table 1). Of the RT-qPCR positive saliva samples collected within a week of symptom onset (n=7), five (71%) and four (57%) were positive with DIAGNOS and Rapid Response, respectively. This was also associated with viral load, with RT-qPCR positive saliva samples collected within one week having a median Ct-value of 25.7 (n=7) compared to a Ct-value of 31.5 for RT-qPCR samples collected after one week (n=8). Nevertheless, it is important to consider that onset of symptoms is based on self-reporting and may not be specifically accurate to COVID-19 onset.

This study has other limitations. One major limitation is that no saliva samples were available from onset of symptoms as i) saliva are not routinely used for SARS-CoV-2 diagnostics and ii) patient recruited in the COVID-19 cohort had been ill for several days before seeking hospital care. Samples were taken in a range of 5-30 days, with a median of 10.5 DPS and thus do not reflect the intended purpose of the RADs, i.e., early after onset of symptoms. Moreover, as we had limited amount of saliva for both the two RADs as well as for the RT-qPCR and cell culture, we had to use less saliva for RNA extraction than is recommended by instructions of the QIAGEN viral extraction kit, thus likely lowering the sensitivity of the RT-qPCR and overestimating the Ct-values. Finally, a relatively low amount of saliva samples was analyzed, thus warranting careful interpretation of the results.

To conclude, using SARS-CoV-2 NCp protein and titrated live SARS-CoV-2, our results show that the rapid saliva antigen test DIAGNOS had similar limit of detection as Panbio, a widely used RAD developed for nasopharyngeal samples. DIAGNOS further exhibited a 47% sensitivity on RT-qPCR positive saliva from COVID-19 hospitalized patients. Sensitivity was however higher on samples collected early after symptom onset, more in line with the intended use of the RADs, corresponding also to a higher viral load. The Rapid Response test showed a higher limit of detection as well as lower sensitivity of RT-qPCR positive saliva samples in hospitalized patients compared to DIAGNOS. The overall results suggest that the DIAGNOS saliva antigen test may be a good and easy-to-use complement and possible self-test to be applied for SARS-CoV-2 detection.

## Data Availability

Data are available upon request

## Acknowledgements

We thank Annette Gustafsson for valuable help with study coordination and patient sampling. We also thank Melissa Govender and Francis Hopkins for help with sample organization and Lennart Svensson for critical reading of the manuscript. This study was supported with a coronavirus ALF grant (30320005) from Region Östergötland, Sweden.

